# Evidence for structural protein damage and membrane lipid remodeling in red blood cells from COVID-19 patients

**DOI:** 10.1101/2020.06.29.20142703

**Authors:** Tiffany Thomas, Davide Stefanoni, Monika Dzieciatkowska, Aaron Issaian, Travis Nemkov, Ryan C. Hill, Richard O Francis, Krystalyn E. Hudson, Paul W. Buehler, James C. Zimring, Eldad A. Hod, Kirk C. Hansen, Steven L. Spitalnik, Angelo D’Alessandro

## Abstract

The SARS-CoV-2 beta coronavirus is the etiological driver of COVID-19 disease, which is primarily characterized by shortness of breath, persistent dry cough, and fever. Because they transport oxygen, red blood cells (RBCs) may play a role in the severity of hypoxemia in COVID-19 patients.

The present study combines state-of-the-art metabolomics, proteomics, and lipidomics approaches to investigate the impact of COVID-19 on RBCs from 23 healthy subjects and 29 molecularly-diagnosed COVID-19 patients. RBCs from COVID-19 patients had increased levels of glycolytic intermediates, accompanied by oxidation and fragmentation of ankyrin, spectrin beta, and the N-terminal cytosolic domain of band 3 (AE1). Significantly altered lipid metabolism was also observed, especially short and medium chain saturated fatty acids, acyl-carnitines, and sphingolipids. Nonetheless, there were no alterations of clinical hematological parameters, such as RBC count, hematocrit, and mean corpuscular hemoglobin concentration, with only minor increases in mean corpuscular volume. Taken together, these results suggest a significant impact of SARS-CoV-2 infection on RBC structural membrane homeostasis at the protein and lipid levels. Increases in RBC glycolytic metabolites are consistent with a theoretically improved capacity of hemoglobin to off-load oxygen as a function of allosteric modulation by high-energy phosphate compounds, perhaps to counteract COVID-19-induced hypoxia. Conversely, because the N-terminus of AE1 stabilizes deoxyhemoglobin and finely tunes oxygen off-loading, RBCs from COVID-19 patients may be incapable of responding to environmental variations in hemoglobin oxygen saturation when traveling from the lungs to peripheral capillaries and, as such, may have a compromised capacity to transport and deliver oxygen.

**Graphical Abstract:** 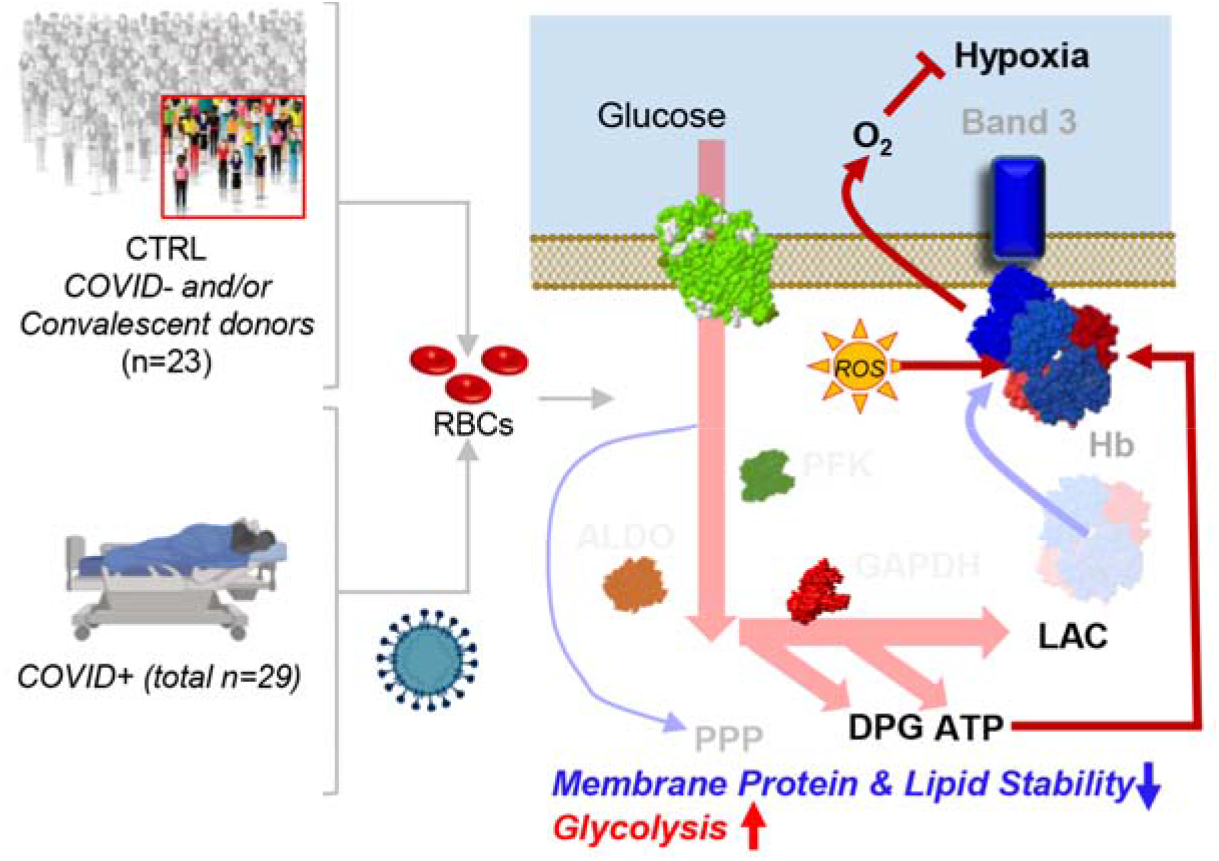

**Key Points:**

- COVID-19 promotes oxidation and fragmentation of membrane proteins, including the N-term of band 3
- RBCs from COVID-19 patients are characterized by increases in glycolysis and altered lipidomes
- COVID-19 impacts two critical mechanisms that finely tune red cell membranes and hemoglobin oxygen affinity

## Introduction

A new RNA coronavirus, SARS-CoV-2, is the etiological agent of a severe acute respiratory syndrome (SARS) and associated complications, collectively termed COronavirus Disease 2019, or COVID-19.^1^ Clinically, COVID-19 is characterized by multiple manifestations, including fever, shortness of breath, persistent dry cough, chills, muscle pain, headache, loss of taste or smell, renal dysfunction, and gastrointestinal symptoms. Analogous to other similar coronaviruses^2^, SARS-CoV-2 penetrates host cells by interactions between its S (spike) protein and the angiotensin converting enzyme receptor 2 (ACE2);^3^ the latter abundantly expressed by lung epithelial cells.^4^ Alternatively, amino acid residues 111-158 of the beta coronavirus S protein can interact with sialic acids on host cell gangliosides, an interaction masked by chloroquines, which were proposed for treating of COVID-19.^5^ Of note, proteomics identified angiotensin and ACE2-interacting proteins on the red blood cell (RBC) surface.^6^ This suggests that RBCs, which cannot support viral replication, may theoretically be invaded by the virus. Indeed, RBCs can be directly or indirectly targeted by pathogens:^7^ infecting pathogens may directly penetrate RBCs (e.g., in malaria), directly promote intravascular hemolysis, or indirectly cause hemolysis or accelerate RBC clearance from the bloodstream by splenic and hepatic reticuloendothelial phagocytes.^7^ Several mechanisms have been proposed to explain these phenomena, including: absorption of immune complexes and complement onto RBC surfaces, development of cross-reacting antibodies, and true autoimmunity with loss of tolerance secondary to infection.^7^ Of note, COVID-19 causes an intense acute phase response and associated complement system dysregulation.^8^

The absence of organelles in mature RBCs resulted in tight physiological regulation, including binding and off-loading oxygen, at the post-translational (e.g., phosphorylation,^9^ methylation^10^) or metabolic level.^11,12^ High-energy phosphate compounds (e.g., 2,3-diphosphoglycerate (DPG), adenosine triphosphate (ATP)) have clear roles in promoting oxygen off-loading.^13^ A recent model proposed that hemoglobin oxygen-saturation and deoxyhemoglobin binding to the cytosolic N-terminus of band 3 (AE1) functions as a sensor of the cell’s redox state and metabolic needs.^14–17^ AE1, the most abundant membrane protein in mature RBCs (∼1 million copies/cell), also participates in the chloride shift (bicarbonate/chloride homeostasis) and as a docking site for several structural proteins critical for membrane integrity.^18^ In this model, high oxygen saturation favors Fenton chemistry in the iron-rich RBC cytosol. In this setting, the AE1 N-terminus is available to bind and inhibit glycolytic enzyme function (i.e., phosphofructokinase (PFK), aldolase (ALDOA), glyceraldehyde 3-phosphate); inhibiting early glycolysis promotes a metabolic shift towards the pentose phosphate pathway (PPP) to generate reducing equivalents (i.e., NADPH) to cope with oxidant stress. In contrast, at low oxygen saturation, deoxyhemoglobin outcompetes the glycolytic enzymes to bind to the AE1 N-terminus, thereby favoring glycolysis and the generation of ATP and DPG to promote further oxygen release and tissue oxygenation, thus relieving hypoxia.^14–17^ Therefore, because RBCs are critical for oxygen transport and off-loading, the severely low oxygen saturations seen in critically ill COVID-19 patients^19^ suggest the importance of determining whether SARS-CoV-2 infection directly and/or indirectly affects RBC metabolism to influence their gas transport, structural integrity, and circulation in the bloodstream.

COVID-19 presents a wide spectrum of signs and symptoms of varying severity; some patients are asymptomatic and others require critical care measures, including ventilation, dialysis, and extracorporeal membrane oxygenation. Disease severity and mortality rates are higher in older males and individuals with other comorbidities, including obesity, diabetes, cardiovascular disease, and immunosuppression (e.g., cancer patients undergoing chemo- or radio-therapy, transplant patients). In contrast, women, children, and adolescents tend to be asymptomatic or mildly symptomatic, while still being contagious and contributing to viral transmission. Of note, age and sex significantly affect RBC metabolism in healthy blood donors with respect to energy and redox metabolism.^20^ As such, we hypothesized that RBC metabolic differences in COVID-19 patients could contribute to their ability to cope with oxidant stress and hypoxemia and, as such, to the heterogeneity of disease expression. In addition to these considerations, preliminary data were offered by others for peer-review supporting a potential direct structural interaction between SARS-CoV-2 proteins and hemoglobins;^19^ if validated, this would provide a direct role for the virus in compromising RBC oxygen transport and delivery.

In light of the above, the present study provides the first comprehensive multi-omics analysis of RBCs from non-infected control and COVID-19 patients, identified by molecular testing of nasopharyngeal swabs.

## Methods

### Blood collection and processing

This observational study was conducted according to the Declaration of Helsinki, in accordance with good clinical practice guidelines, and approved by the Columbia University Institutional Review Board. Subjects seen at Columbia University Irving Medical Center/New York-Presbyterian Hospital included 29 COVID-19-positive patients, as determined by SARS-CoV-2 molecular testing of nasopharyngeal swabs. The control group included 23 subjects, all of whom were molecularly SARS-CoV-2 negative by nasopharyngeal swab at the time of the blood draw. Some patients in the control group were “never positive” subjects and some were COVID-19 convalescent patients, and potential convalescent plasma donors, who were previously positive, but currently negative, as determined by testing nasopharyngeal swabs, and at least 14 days post-resolution of symptoms. RBCs obtained by centrifugation (1,500 x *g* for 10 min at 4°C) of freshly drawn blood samples were collected and then de-identified. RBCs were extracted via a modified Folch method (chloroform/methanol/water 8:4:3 *v/v/v*), which completely inactivates other coronaviruses, such as MERS-CoV ^21^. Briefly, RBC pellets (20 μL) were diluted in 130 μl of LC-MS grade water, 600 μl of ice-cold chloroform/methanol (2:1) was added; wthe samples were vortexed for 10 seconds, incubated at 4°C for 5 minutes, quickly vortexed (5 seconds), and centrifuged at 14,000 x *g* for 10 minutes at 4°C. The top (i.e., aqueous) phase was transferred to a new tube for metabolomics, the bottom phase for lipidomics, and the interphase protein disk for proteomics. In a biosafety hood, the protein disk was rinsed with methanol (200 ul) before centrifugation (14,000 x *g* for 4 minutes) and subsequent air drying.

### Protein digestio

Protein pellets from RBC samples were digested in an S-Trap filter (Protifi, Huntington, NY), following the manufacturer’s procedure. Briefly, ∼50 μg of protein were first mixed with 5% SDS. Samples were reduced with 10 mM dithiothreitol at 55°C for 30 minutes, cooled to room temperature, and then alkylated with 25 mM iodoacetamide in the dark for 30 minutes. Phosphoric acid was then added to a final concentration of 1.2% followed by 6 volumes of binding buffer (90% methanol; 100 mM triethylammonium bicarbonate (TEAB); pH 7.1). After gentle mixing, the protein solution was loaded onto an S-Trap filter, centrifuged (2000 x *g*; 1 minute), and the flow-through collected and reloaded onto the filter. This step was repeated three times, and then the filter was washed with 200 μL of binding buffer 3 times. Finally, 1 μg of sequencing-grade trypsin and 150 μL of digestion buffer (50 mM TEAB) were added onto the filter and digested at 47°C for 1 hour. To elute peptides, three step-wise buffers were applied, with 200 μL of each with one more repeat; these included 50 mM TEAB, 0.2% formic acid in water, and 50% acetonitrile and 0.2% formic acid in water. The peptide solutions were pooled, lyophilized, and resuspended in 0.1 % formic acid.

### Nano Ultra-High-Pressure Liquid Chromatography-Tandem Mass Spectrometry (MS) proteomics

Samples (200 ng each) were loaded onto individual Evotips for desalting and then washed with 20 μL 0.1% formic acid followed by adding 100 μL of storage solvent (0.1% formic acid) to keep the Evotips wet until analysis. The Evosep One system was coupled to the timsTOF Pro mass spectrometer (Bruker Daltonics, Bremen, Germany). Data were collected over an m/z range of 100-1700 for MS and MS/MS on the timsTOF Pro instrument using an accumulation and ramp time of 100 milliseconds. Post processing was performed with PEAKS studio (Version X+, Bioinformatics Solutions Inc., Waterloo, ON). Pathway analyses were performed with DAVID software and Ingenuity Pathway Analysis. Graphs and statistical analyses were prepared with GraphPad Prism 8.0 (GraphPad Software, Inc, La Jolla, CA), GENE E (Broad Institute, Cambridge, MA, USA), and MetaboAnalyst 4.0.^22^

### Ultra-High-Pressure Liquid Chromatography-Mass Spectrometry (MS) metabolomics and lipidomics

Metabolomics and lipidomics analyses were performed using a Vanquish UHPLC coupled online to a Q Exactive mass spectrometer (ThermoFisher, Bremen, Germany). Samples were analyzed using 5, 15, and 17 min gradients, as described ^23,24^. For targeted quantitative experiments, extraction solutions were supplemented with stable isotope-labeled standards, and endogenous metabolite concentrations were quantified against the areas calculated for heavy isotopologues for each internal standard ^23,24^. Data were analyzed using Maven (Princeton University) and Compound Discoverer 2.1 (ThermoFisher). Graphs and statistical analyses were prepared with GraphPad Prism 8.0, GENE E, and MetaboAnalyst 4.0.^25^ Spearman’s correlations and related p-values were calculated with R Studio.

## Results

### COVID-19 influences RBC metabolism and proteome

Metabolomics and proteomics analyses were performed on red blood cells (RBCs) from COVID-19 negative (n=23) and positive (n=29) subjects (**Figure 1.A**). With the exception of minor increases in mean corpuscular volume (MCV), standard hematological parameters did not significantly differ between the two groups, including RBC count, hematocrit (HCT), hemoglobin (Hgb), mean corpuscular hemoglobin (MCH), mean corpuscular hemoglobin concentration (MCHC), and RBC distribution width (RDW; **Supplementary Figure 1**). Targeted metabolomics and proteomics analyses (**Supplementary Table 1**) identified COVID-19 effects on RBCs, as gleaned by partial least square discriminant analysis (PLS-DA; **Figure 1.B**) and hierarchical clustering analysis of the top 50 significant metabolites (**Figure 1.C**) and proteins (**Figure 1.D**) sorted by t-test. A vectorial version of these figures is provided in **Supplementary Figures 2** and **3**, respectively. Volcano plot analyses identified significant RBC proteins and metabolites when comparing COVID-19 positive and negative subjects (**Figure 1.E**); similar analyses were performed using untargeted metabolomics data (**Supplementary Figure 4**). Pathway analyses based on these results (**Figure 1.F**) highlighted a significant effect of COVID-19 on protein degradation pathways (including proteasome and ubiquitinylation/NEDDylation components), ferroptosis, cyclic-AMP and AMPK signaling cascades, and lipid metabolism (especially acyl-carnitines and sphingolipid metabolism; **Figure 1.F**).

**Figure 1.**
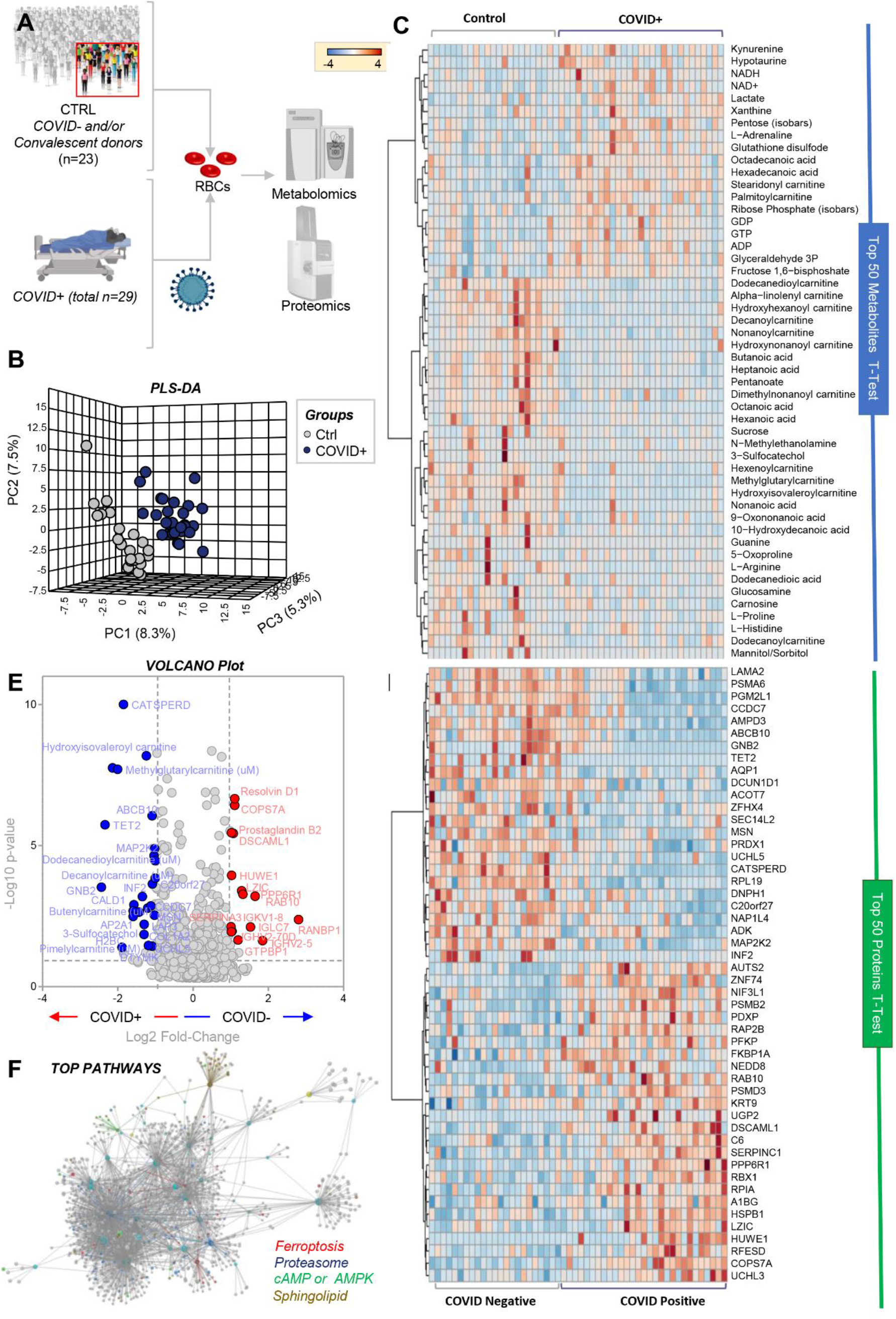
RBC metabolism and proteome are influenced by COVID-19. Metabolomics and proteomics analyses were performed on RBCs from COVID-19-negative (n=23) and -positive (n=29) subjects, as determined by molecular testing of nasopharyngeal swabs (**A**). The effects of COVID-19 on RBCs, as gleaned by PLS-DA (**B**) and hierarchical clustering analysis of the top 50 metabolites (**C**) and proteins (**D**) by t-test. In **E**, the volcano plot highlights the significant metabolites and proteins increasing (red) or decreasing (blue) in RBCs from COVID-19 patients, as compared to non-infected controls. In **F**, pathway analyses were performed on the significant features from the analyses in **B-E**.

### Energy and redox metabolism in RBCs from COVID-19 positive patients

RBCs from COVID-19 patients had significant alterations in glycolysis (**Figure 2.A**). Specifically, they exhibited significant increases in sucrose consumption and accumulation of several glycolytic intermediates, as compared to controls, including glucose 6-phosphate, fructose bisphosphate, glyceraldehyde 3-phosphate, 2,3-diphosphoglycerate (*p-value = 0*.*075*), phosphoglycerate, phosphoenolpyruvate, pyruvate, lactate, and NADH (**Figure 2.A**). This phenomenon was explained, at least in part, by the apparent higher levels of phosphofructokinase (PFK), the rate limiting enzyme of glycolysis, in RBCs from COVID-19 subjects as compared to controls. There were also significant decreases in levels of phosphoglucomutase 2-like 1 (PGM2L1), which catalyzes the synthesis of hexose bisphosphate and, thus, slows down glycolysis, and glyceraldehyde 3-phosphate dehydrogenase (GAPDH), a redox sensitive enzyme that limits flux through late glycolysis.^17^ In contrast, ribose phosphate (isobars), the end product of the PPP, significantly accumulated in RBCs from COVID-19 patients, suggesting greater oxidant stress in these RBCs (**Figure 2.B**). Consistently, RBCs from COVID-19 patients had increased oxidized glutathione (GSSG), but not reduced glutathione (GSH); correspondingly, decreases were seen in 5-oxoproline, a metabolic end product of the RBC gamma-glutamyl cycle (**Figure 2.C**). In contrast, RBCs from COVID-19 patients had higher levels of carboxylic acids (alpha-ketoglutarate, fumarate; **Figure 3.A**), and higher levels of total adenylate pools (ATP, ADP, AMP; **Figure 3.A**). Purine deamination and oxidation products were not significantly increased, with the exception of xanthine; however, significantly lower levels of enzymes involved in purine metabolism were observed in RBCs from COVID-19 patients, specifically AMP deaminase 3 (AMPD3) and adenylate kinase (ADK; **Figure 3.A**).

**Figure 2.**
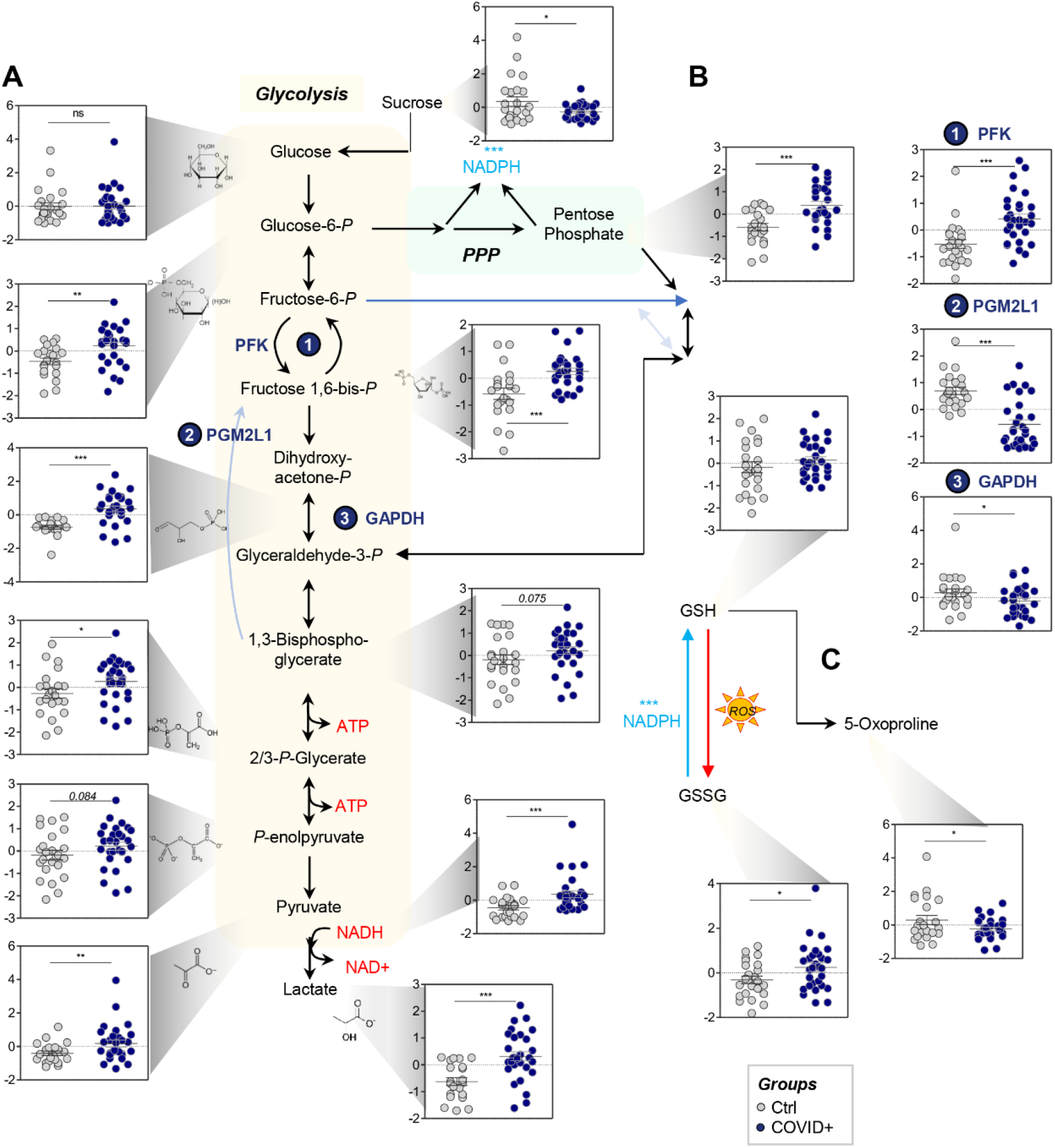
COVID-19 significantly affects RBC glycolysis (A) and the pentose phosphate pathway (PPP - B), with no significant effect on glutathione homeostasis (C). Metabolomics of RBCs from COVID-19 subjects identified significant increase in several glycolytic intermediates, as compared to controls, including glucose 6-phosphate, fructose bisphosphate, glyceraldehyde 3-phosphate, 2,3-diphosphoglycerate, lactate, and NADH. This phenomenon was at least in part explained by the higher protein levels of PFK, the rate limiting enzyme of glycolysis, in RBCs from COVID-19 subjects, as compared to controls. These subjects also had significant decreases in the levels of PGM2L1, which catalyzes the synthesis of hexose bisphosphate and, thus, slows down glycolysis, and GAPDH, a redox-sensitive enzyme. On the other hand, ribose phosphate (isobars), the end product of the PPP, significantly accumulated in RBCs from COVID-19 patients, suggesting a higher degree of oxidant stress in these RBCs; this was confirmed, in part, by significantly higher levels of GSSG and lower levels of 5-oxoproline (C). Asterisks indicate significance by t-test (* *p<0*.*05; ** p<0*.*01; *** p<0*.*001*). Groups are color coded according to the legend in the bottom right corner of the figure.

**Figure 3.**
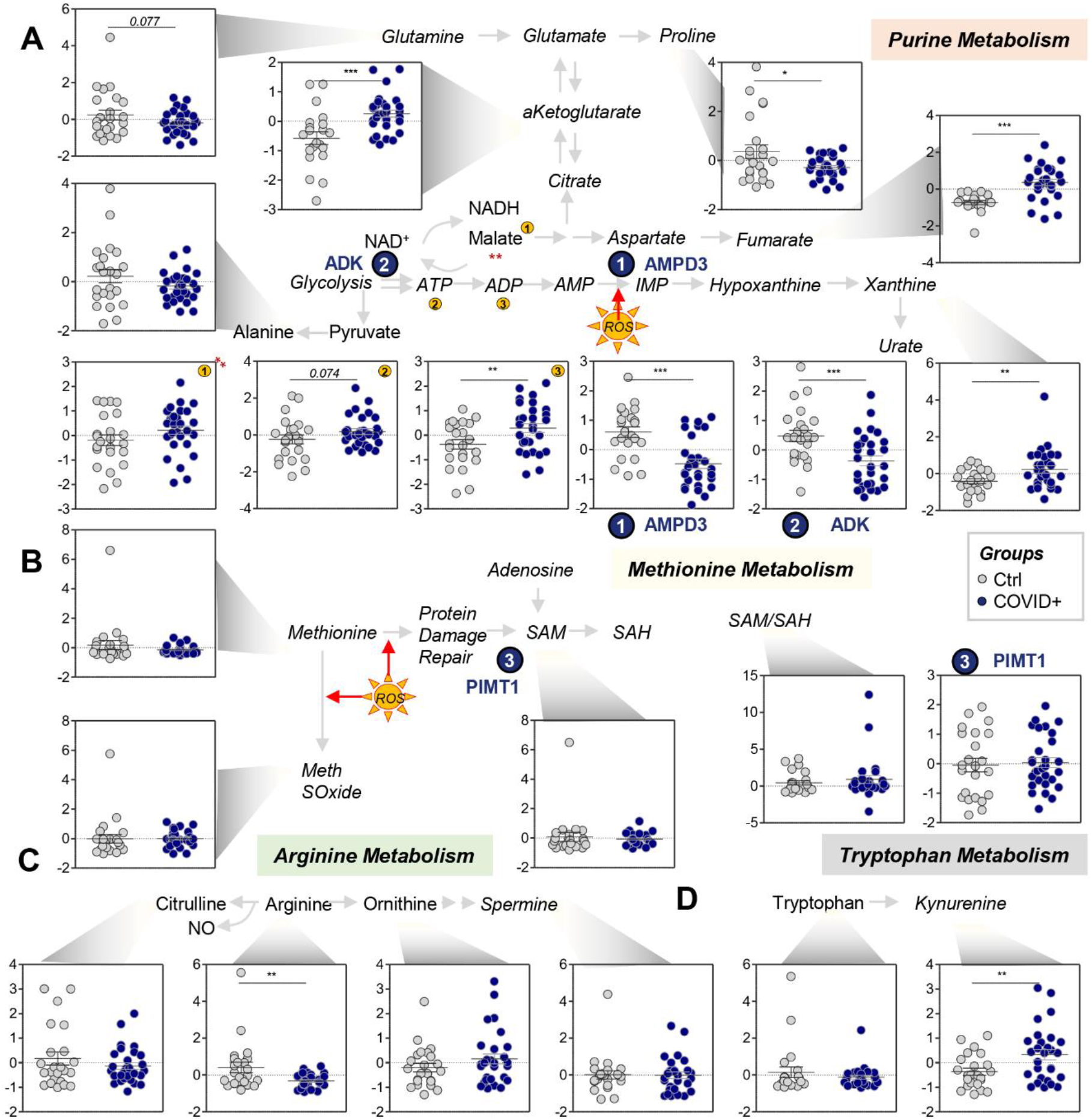
COVID-19 significantly affects transamination and carboxylic acid metabolism in RBCs (A), but not purine deamination (B), with only limited effects on arginine (C) and tryptophan (D) metabolism. Asterisks indicate significance by t-test (* *p<0*.*05; ** p<0*.*01; *** p<0*.*001*). Groups are color coded according to the legend in the center of the figure.

No significant alterations were observed for methionine levels, consumption (e.g., to generate S-adenosyl-methionine for isoaspartyl damage-repair by PIMT1), or oxidation (i.e., methionine sulfoxide; **Figure 3.B**). However, significantly lower arginine levels were accompanied by (non-significant) trends towards increased and decreased levels of ornithine and citrulline, respectively, suggesting potentially increased arginase, and decreased nitric oxide synthase, activity in RBCs from COVID-19 patients (**Figure 3.C**). In contrast, increased tryptophan oxidation to kynurenine was observed in RBCs from COVID-19 patients, in the absence of alterations in tryptophan levels (**Figure 3.D**).

In the light of this apparent oxidant stress-related signature, we hypothesized that RBCs from COVID-19 patients may suffer from impaired antioxidant enzyme machinery, perhaps triggered by degradation of redox enzymes in the context of ablated *de novo* protein synthesis capacity in mature RBCs. Although enzyme levels do not necessarily predict enzymatic activity, relative quantities of the main antioxidant enzymes are plotted in **Figure 4.A**, including catalase (CAT), peroxiredoxins (PRDX) 1, 2, and 6, glutathione peroxidases (GPX) 1 and 4, superoxide dismutase (SOD1), gamma-glutamyl cysteine ligase (GCLC), glutathione reductase (GSR), glucose 6-phosphate dehydrogenase (G6PD), and biliverdin reductase B (BLVRB). Notably, PRDX1, SOD1, and G6PD were significantly decreased (**Figure 4.A**), suggesting possible degradation of these enzymes in RBCs from COVID-19 patients. Indeed, these RBCs had higher levels of the components of the proteasome and degradation machinery, such as the ubiquitin-like protein NEDD8, cullin-associated NEDD8-dissociated protein 1 (CAND1), E3 ubiquitin-protein ligase HUWE1, along with decreases in proteasomal subunit A6 (PSMA6), a part of the ATP-dependent 25S proteasome (**Figure 4.B**). Taken together, these results suggest increased RBC protein degradation in COVID-19.

**Figure 4.**
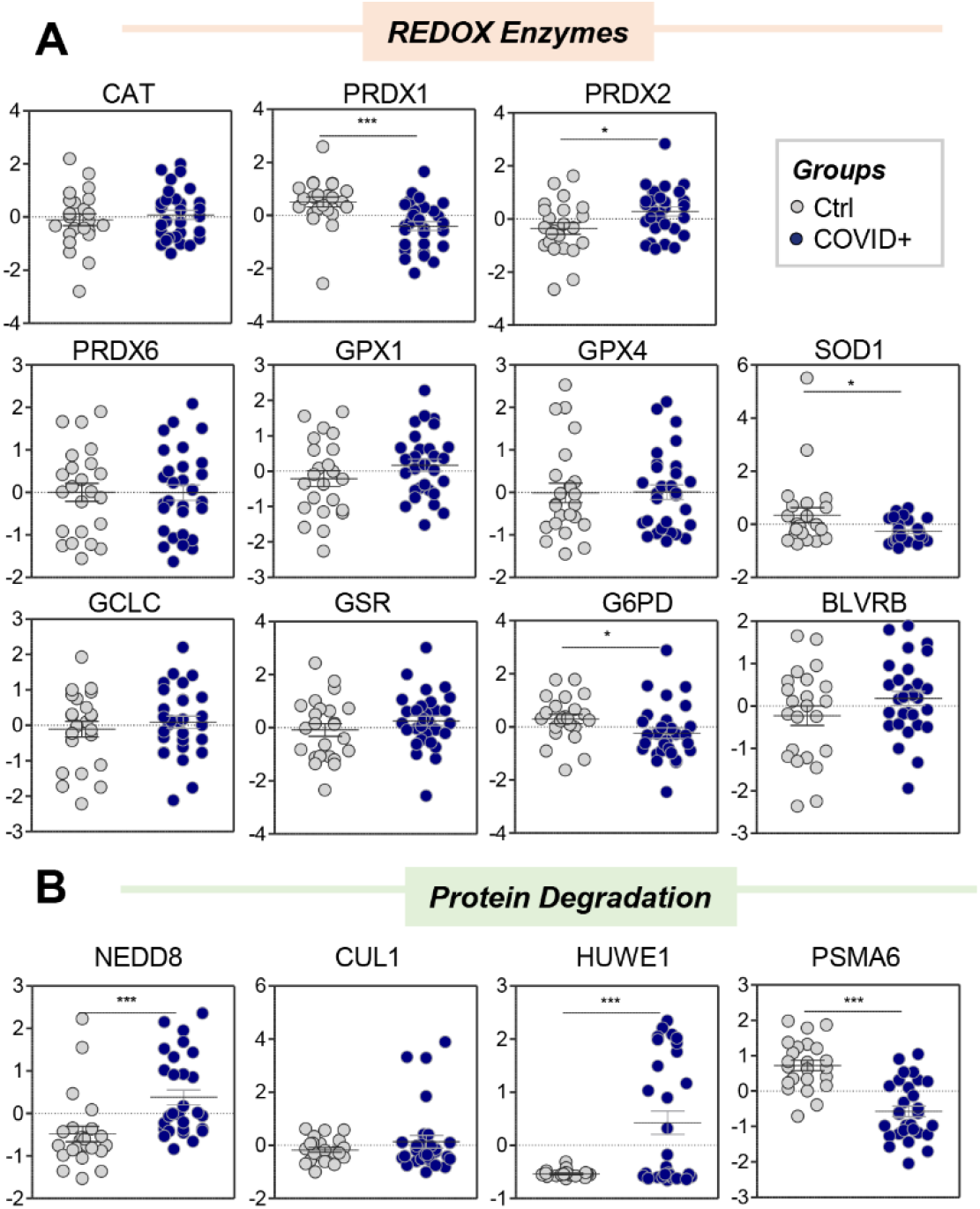
RBCs from COVID-19 patients have limited alterations in antioxidant enzyme levels (A), but increased levels of components of the ubiquitination/NEDDylation system (B). Asterisks indicate significance by t-test (* *p<0*.*05; ** p<0*.*01; *** p<0*.*001*). Groups are color coded according to the legend in the bottom right corner of the figure.

### COVID-19 influences oxidation and structural integrity of key RBC proteins

Despite no significant changes in the total levels of key structural proteins (e.g., spectrin alpha (SPTA1) and ankyrin (ANK1); **Figure 5.A**), proteomics analysis showed minor increases in AE1 in RBCs from COVID-19 patients. To explain this observation, we hypothesized that increases in AE1 solubility and detection via proteomics approaches could be, at least in part, explained by protein fragmentation secondary to (oxidant) stress in these patients (**Supplementary Figure 1**). To explore the hypothesis, peptidomics analyses were performed, identifying significant increases/decreases in specific peptides from most structural proteins (heat map in **Figure 5.B** shows the top 50 significant changes by t-test). Further analysis of AE1-specific peptides from RBCs from COVID-19 patients highlighted significantly increased levels of AE1 peptides spanning amino acid residues 57-74, along with decreased levels from N-terminal residues 1-57 (**Figure 5.C**), as mapped (red) against the pdb:1hyn spanning residues 56-346 of AE1 (grey; **Figure 5.D**). In addition, COVID-19 patient RBC AE1 was significantly more oxidized (determined by the cumulative peak area of peptide hits carrying redox modifications: M oxidation + N/Q deamidation), as compared to controls (**Figure 5.E**), in the absence of detectable changes in peptide levels beyond residue 75 (**Figure 5.F**). Similar increases in the levels and oxidation of peptides from SPTA1 (**Figure 5.G**) and ANK1 (**Figure 5.H**) were observed, consistent with an apparent effect of COVID-19 on the structural integrity of RBC membrane proteins (**Figure 5.I**).

**Figure 5.**
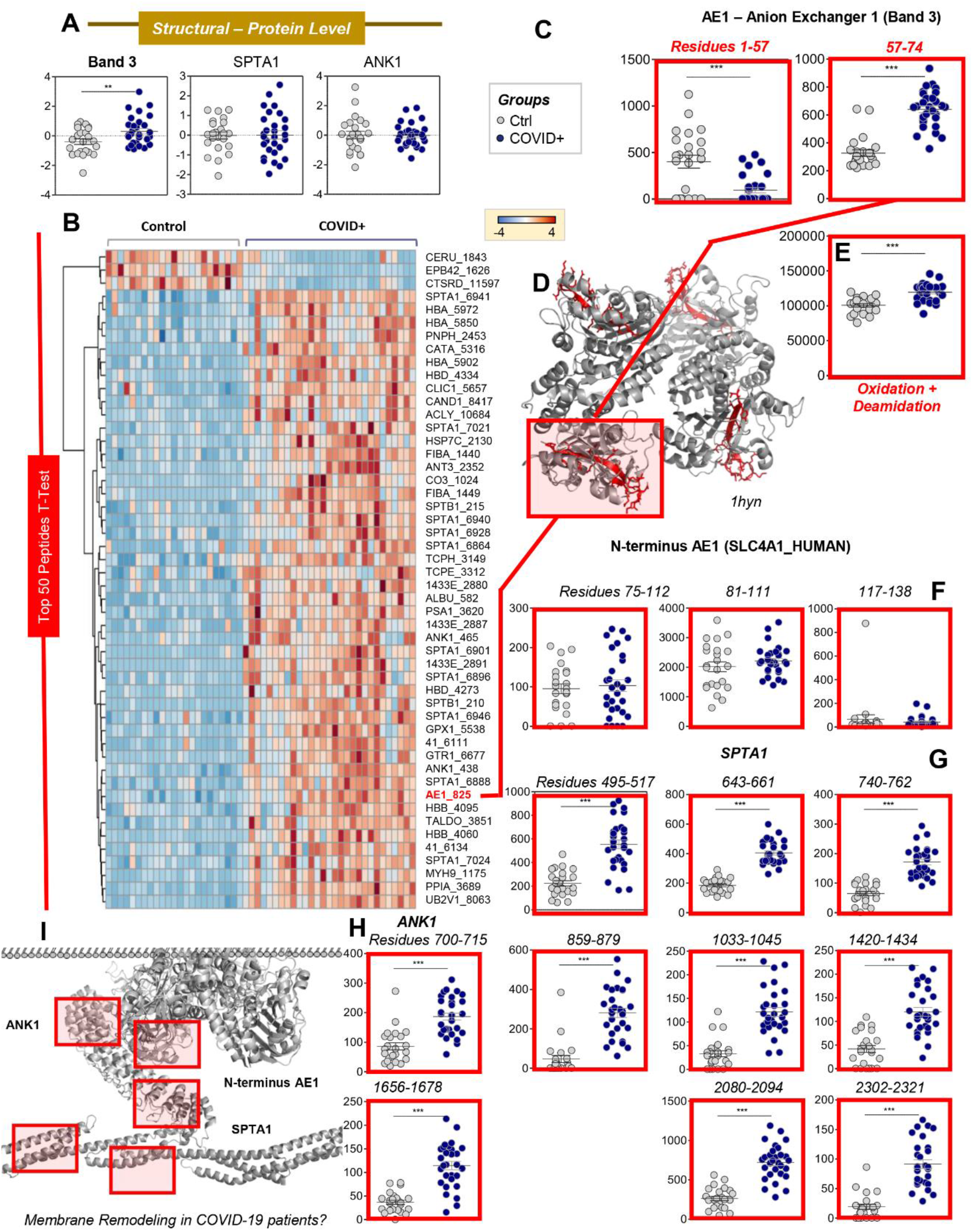
COVID-19 promotes oxidation and alteration of key structural RBC proteins. Despite no significant changes in the total levels of key structural proteins (e.g., band 3: AE1; spectrin alpha: SPTA1; ankyrin: ANK1; **A**), peptidomics analyses showed significant increases and decreases in specific peptides from these proteins (heat map in **B** shows the top 50 significant changes by t-test). Further analysis of AE1 identified significant increases in levels of the peptide spanning amino acid residues 57-74, in contrast to decreased levels of the N-terminal 1-57 peptide (**C**), as mapped (red) against the pdb:1hyn spanning residues 56-346 of AE2 (grey; **D**). In addition, in COVID-19 patients, RBC AE1 was significantly more oxidized (M oxidation + N/Q deamidation) than in control RBCs (**E**), in the absence of detectable changes in the levels of peptides beyond residue 75 (**F**). Similar increases in the levels and oxidation of peptides for SPTA1 (**G**) and ANK1 (**H**) were observed, consistent with an effect of COVID-19 on the integrity of RBC structural membrane proteins (**I**).

### RBCs from COVID-19 patients exhibit significantly altered membrane lipids and lipid remodeling pathways

Lipidomics analyses also suggested alterations in RBC membrane integrity in COVID-19 patients. First, despite comparable RDW and slightly increased MCV (**Supplementary Figure 1**), RBCs from COVID-19 patients had significantly lower levels of short- and medium-chain acylcarnitines (i.e., C5-OH, C6-OH, C6:1, C9, C9-OH, C9-dME, C10, C12, C12:2; **Figure 6.A**), but not with long-chain fatty acyls groups (C16, C18), unless unsaturated (18:3l **Figure 6.A**). These were accompanied by decreased short-chain fatty acids (C4:0, C5:0, C6:0, C7:0, C8:0, C9:0 C10:0-OH) and increased long-chain saturated fatty acids (C16:0 and C18:0 (palmitate and stearate, respectively); **Figure 6.B**), with no detectable changes in mono- or poly-unsaturated fatty acid levels (**Supplementary Figure 5**). RBCs from COVID-19 patients also had increased oxylipin derivatives of unsaturated fatty acids, including 5-oxoETE, 13-HODE, and resolvins D1, D2, E1 (**Figure 6.C**).

**Figure 6.**
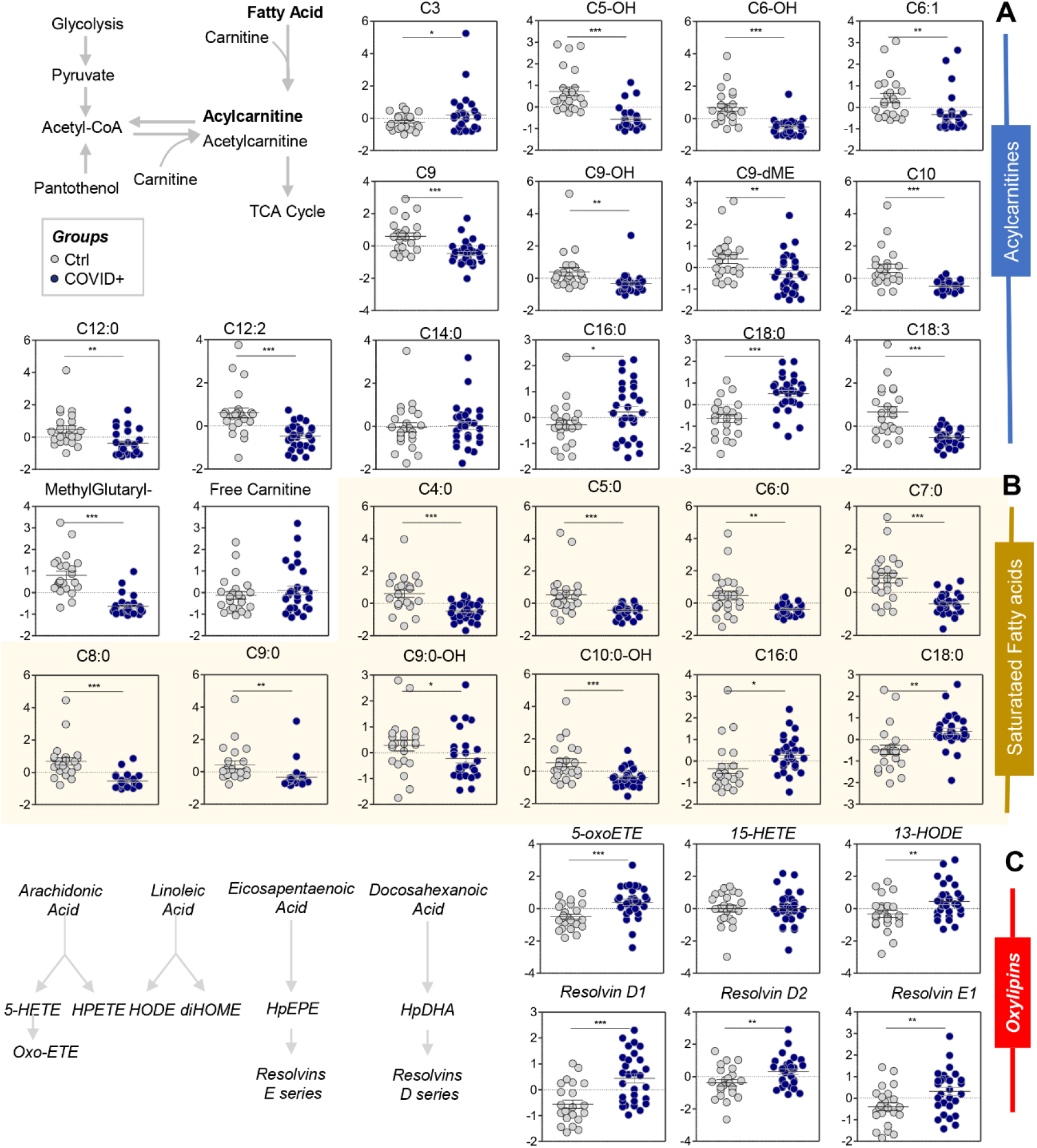
RBC acyl-carnitines (A), saturated fatty acids (B), and oxylipins and resolvins (C) were significantly affected by COVID-19. Asterisks indicate significance by t-test (* *p<0*.*05; ** p<0*.*01; *** p<0*.*001*). Groups are color coded according to the legend in the top left corner of the figure.

To determine whether the observed changes in free fatty acid and acyl-carnitine levels were driven by a specific class of lipids, in-depth lipidomics analyses of RBCs from COVID-19 negative and positive patients were performed (**Figure 7; Supplementary Table 1**). Of all the lipid classes investigated, there were significant alterations in levels of phosphatidic acids (PAs), sterols (ST), sphingolipids (SPH), and lysophosphatidic acids (LPA) (log10 normalized areas are shown in **Figure 7.A**). Volcano and bar plot representations of the most significantly affected lipid classes or lipids are shown in **Figure 7.B-D**. This analysis identified significant decreases in SPH, CmEs, LPAs, and cPAs, and increases in ceramide-phosphorylethanolamine (CerPE), as the most affected classes in COVID-19 patients. In addition, although most lipid classes decreased significantly in COVID-19 patients, several phosphatidylethanolamines (PEs) increased significantly, including PE 30:3, 36:2, and 37:2. Likewise, despite an overall trend towards decreases in phosphatidylcholines (PCs), some specific PCs significantly increased in COVID-19 RBCs (e.g., 34:2; **Figure 7.D**).

**Figure 7.**
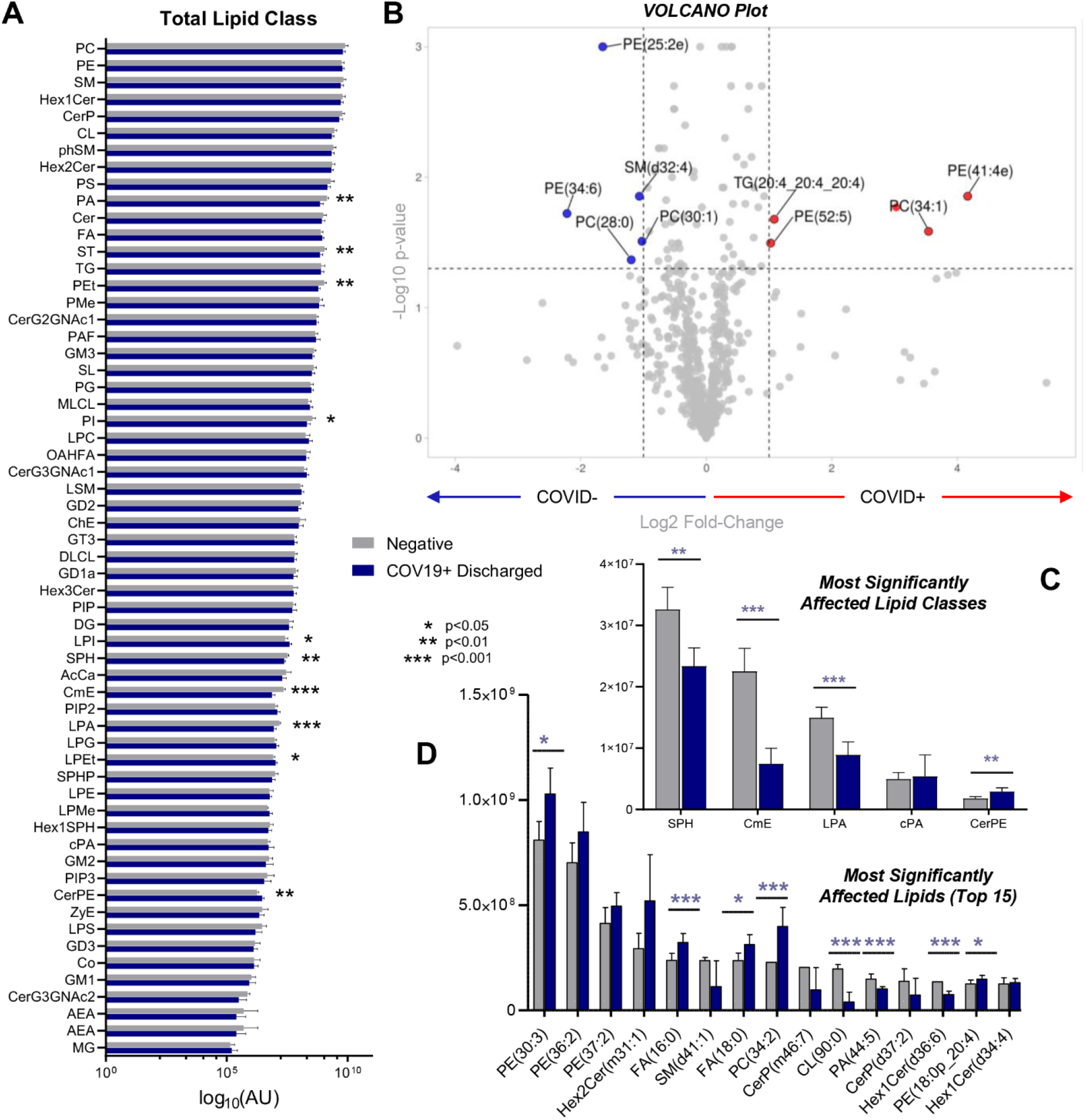
Lipidomics analyses of RBCs from COVID-19-positive and control patients. (**A**) An overview of all the lipid classes investigated in this study (log10 normalized areas) and their variation across groups (symbols indicating significance by t-test are reported in the legend in the center of the panel). (**B**) Volcano plot showing the most significantly affected lipids, comparing COVID-19-positive subjects and controls. (**C** and **D)** Expanded view of the top lipid classes and lipids, respectively, affected by COVID-19. Asterisks indicate significance by t-test (* *p<0*.*05; ** p<0*.*01; *** p<0*.*001*). Groups are color coded according to the legend in the center of the figure.

## Discussion

The present study provides the first multi-omics characterization of RBCs from COVID-19 patients. We identified increased glycolysis in RBCs from COVID-19 patients, accompanied by increased oxidation (deamidation of N, oxidation of M, methylation of D,E) of key structural proteins, including the N-terminus of AE1, ANK1, and SPTA1. These changes were accompanied by lower levels of acyl-carnitines, free fatty acids, and most lipids (especially sphingolipids, phosphatidic acids, and phosphatidylethanolamines), despite minor increases in MCV, and in the absence of significant changes in RBCs count, HCT, or other clinical hematological parameters. Interestingly, fragmentation/oxidation of the N-terminus of AE1 is expected to disrupt the inhibitory binding of glycolytic enzymes, thereby promoting flux through glycolysis; in turn, hemoglobin oxygen off-loading would be favored via allosteric modulation by RBC DPG (increased in COVID-19) and ATP (trend towards increase) to counteract hypoxia; this interpretation reconciles the metabolomics and peptidomics findings in this study. Conversely, one can speculate that, similar to what is observed with genetic variants that favor the splicing of N-terminal amino acids 1-11 of AE1 (i.e., band 3 Neapolis^26^), RBCs from COVID-19 subjects may have increased susceptibility to oxidant stress-induced lysis and impaired ability to off-load oxygen, because their AE1 would be less able to bind and (i) inhibit glycolytic enzymes, rediverting metabolic fluxes to the PPP to generate reducing equivalents; (ii) stabilize the tense, deoxygenated state of hemoglobin.^27,28^ Unfortunately, owing to logistical limitations, we could not directly measure RBC parameters directly related to gas transport physiology, a limitation that we will address in follow-up studies.

It is not clear whether the alterations of the N-terminus of AE1 are driven by oxidant stress alone or by an enzymatic activity secondary to the infection (e.g., calcium-activated proteases). However, although SARS-CoV-2 encodes cleaving enzymes (e.g., papain-like proteases), comparing our proteomics data with this viral genome did not produce any positive identifications.

This suggests that the virus does not penetrate RBCs or, if it does, its protein components are rapidly degraded and not resynthesized owing to the lack of organelles; alternatively, our approach may not be sensitive enough to detect trace viral proteins in the background of ∼250 million hemoglobin molecules per RBC.^29^

Modification and oxidation of the N-termini of AE1, ANK1, and SPTB were accompanied by altered acyl-carnitines, fatty acids (especially saturated short- and medium-chain fatty acids), and lipid metabolism (especially sphingolipids). The latter is interesting because signaling through the N-terminus of AE1 mechanistically cross-regulates with sphingolipids to promote hemoglobin oxygen off-loading in response to physiological (e.g., high-altitude) or pathological (e.g., sickle cell disease) hypoxia.^15,30^ This signature is consistent with impaired membrane lipid homeostasis, which is not attributable to ATP depletion (not significantly altered in COVID-19 patients). Interestingly, viral infection,^31,32^ including SARS-CoV-2,^33^ is associated with altered fatty acid and acyl-carnitine profiles secondary to phospholipase A2 activation. Of note, a redox sensitive enzyme that is abundant in RBCs, peroxiredoxin 6, also exerts phospholipase A2-like activity.^34^ Although PRDX6 levels did not significantly differ between COVID-19 patients and controls, it is interesting that several classes of lysophospholipids were altered in COVID-19 patients. As such, because of the large number of circulating RBCs (∼25 trillion in an adult), one may speculate that the increases in serum fatty acids in COVID-19 patients^33^ may, at least in part, be due to decreases in the same fatty acids in the erythroid compartment. These fatty acids are critical building blocks that sustain proliferation of replicating viruses, to the extent that they support viral membrane formation prior to decoration with nucleocapsid and spike proteins, as the virus is assembled in target cells^31,32^ (i.e., not RBCs).

Although data regarding disease severity were not available for the subjects in this study, one common manifestation of COVID-19 is persistent high fever. Interestingly, RBCs exhibit increased vesiculation and altered acyl-carnitine metabolism in response to severe increases in temperature *in vivo* and *in vitro*;^35,36^ ATP-depletion and/or activation of the Lands cycle^35,37^ were proposed as candidate mechanisms to explain these findings.

We also identified increases in carboxylic acids and pentose phosphate isobars. In other settings (e.g., the iatrogenic interventions of refrigerated blood storage for clinical purposes^38^ or heat shock^35^), RBC accumulation of these metabolites was consistent with increased oxidant stress-dependent AMPD3 catabolism of high-energy purines. However, with the exception of xanthine, increases in oxidized purines were not observed in COVID-19 patient RBCs, despite higher levels of AMPD3 and lower levels of ADK in these RBCs. In contrast, there were increased steady-state levels of ribose phosphate (and pentose phosphate isobars), a marker of PPP activation in response to oxidant stress in RBCs,^17^ despite decreased levels of G6PD, the rate limiting enzyme of this pathway. Although relative levels may not reflect enzymatic activity, one may speculate that the effects of COVID-19 on RBC biology may be exacerbated when the stability and activity of G6PD are modified by natural mutations. Thus, G6PD deficiency^20,39^ is the most common human enzymopathy, affecting ∼400 million people world-wide; it also disproportionally affects particular ethnic groups, including African Americans, who are more susceptible to developing severe COVID-19. Because G6PD is also an X-linked gene, it may also partially explain the sex-dependent component of COVID-19 severity, with worse outcomes in male patients.^40^ However, despite 14 male and 9 female subjects in the control group, and 18 males and 11 females in the COVID-19 group, the present study did not identify major sex-specific signatures for COVID-19 RBCs. Despite the oxidant stress signature observed in COVID-19 RBCs, there were no increases in methionine consumption or oxidation, hallmarks of isoaspartyl-damage repair of RBC proteins following oxidant insults.^10^ Conversely, COVID-19 RBCs exhibited decreased levels of key antioxidant enzymes (PRDX1, SOD1, G6PD) and increased markers of protein degradation (e.g., via the ubiquitinylation-proteasome system). PRDX2 was a notable exception; increased levels of this protein may be due to it increased solubility when released from the membrane, where binds to the N-terminus of AE1,^41^ which was damaged in COVID-19 RBCs. Increased oxidation of structural proteins, along with alterations of lipid compartments, may alter RBC deformability following SARS-CoV-2 infection. Importantly, the role of RBC morphology and deformability in clot formation and stability are increasingly appreciated.^42,43^ These RBC parameters are tightly regulated by structural protein homeostasis and by the availability of high-energy phosphate compounds required to maintain ion and structural lipid homeostasis (e.g., membrane exposure of phosphatidylserine).^35,44^ As such, the altered RBC structural proteins in COVID-19 may contribute to the thromboembolic and coagulopathic complications seen in some critically ill patients; nonetheless, larger studies will be necessary to test this hypothesis.

Increased levels of kynurenine in RBCs from COVID-19 patients were consistent with prior observations in sera.^33^ Although this is likely due to equilibrium between kynurenine levels in RBCs and the extracellular environment, it is interesting that increased levels of kynurenine were observed in male, but not female, RBCs following storage-induced oxidant stress of leukocyte- and platelet-reduced RBC concentrates.^20,45^

ABO blood type may be associated with COVID-19 disease severity. In preliminary studies, COVID-19 incidence and severity were increased in Group A subjects, whereas Group O subjects were less affected^46,47^. However, the present study was insufficiently powered to determine the impact of blood type on COVID-19-induced effects on the RBC metabolome and proteome.

Additional limitations of this study pertain to the lack of clinical information on disease severity and stage for the studied COVID-19 patients, a limitation that we will address with currently ongoing prospective enrollment of patients for future study. Similarly, the present study was not sufficiently powered to determine the impact of COVID-19 on RBCs as a function of other biological variables, including subject sex, age, ethnicity, blood type, and habits (e.g., smoking); these are all associated with the RBC’s capacity to cope with oxidant stress and modulate energy metabolism.^20,48–52^

## Data Availability

All the raw data for this manuscript are provided in Supplementary Table 1

## Acknowledgments

This research was supported by funds from the Boettcher Webb-Waring Investigator Award (ADA), RM1GM131968 (ADA and KCH) from the National Institute of General and Medical Sciences, and R01HL146442 (ADA), R01HL149714 (ADA), R01HL148151 (ADA, SLS, JCZ), R21HL150032 (ADA), and T32 HL007171 (TN) from the National Heart, Lung, and Blood Institute.

## Authors’ contributions

TT, ROF, SLS, EAH designed the study. TT, ROF, EAH collected and processed the samples. DS, TN, MD, RCH, ADA performed omics analyses. ADA performed data analysis and prepared figures and tables. ADA wrote the first draft of the manuscript, which was significantly revised by SLS, TT, PWB, JCZ and KEH, and finally approved by all the authors.

